# You can’t manage what you can’t imagine: The Digital Health Checklist-Risk Management (DHC-RM) Tool to enhance participant protections in digital health research

**DOI:** 10.64898/2026.02.22.26346854

**Authors:** Alan J. Card, Daniela Vital, Camille Nebeker

## Abstract

Digital health technologies are powerful–enhancing data collection, participant engagement, and personalized health interventions–yet their rapid proliferation has outpaced guidance for research participant protection. Current practice assists researchers in identifying risks but provides limited support for comprehensive risk management. To address this gap, we developed the Digital Health Checklist–Risk Management (DHC-RM) Tool, which integrates the established Digital Health Checklist with approaches from safety risk management.

We conducted a study (n=40) comparing the DHC-RM Tool with current practice using a randomized experimental difference-in-differences design. Primary outcomes were the quantity, variety, and novelty of risks identified; secondary outcomes were the same constructs applied to risk control development.

Compared with current practice, use of the DHC-RM Tool resulted in dramatically improved performance across all primary outcomes. Users identified on average 14.7 additional risks (compared to baseline) versus 0.26 in the control group and a higher number of risks in each of six pre-identified risk domains. Half of all distinct risks identified in the comparison phase were identified exclusively using the tool. The tool also improved risk control design, producing 9.63 additional risk control strategies per participant compared with 0.15 for current practice and yielding substantially greater novelty and variety.

User feedback was also positive: 75% of participants reported they would use the tool again, citing its structured workflow, just-in-time examples, improved insight into risks, and its value for IRB communication. Suggestions for refinement focused primarily on expanding training examples and providing additional support for risk control development.

The DHC-RM Tool significantly improves risk management practice in digital health research. By embedding structured, ethics-informed risk management into digital health research design, the DHC-RM Tool has the potential to improve participant protection while also streamlining ethics approval.

**Author Summary:** Digital health research can put participants (and others) at risk in ways that don’t always occur to the researchers who are designing a study. Researchers also face challenges in prioritizing risks and coming up with ideas to reduce those risks. We developed a new approach, *the Digital Health Checklist – Risk Management Tool (DHC-RM Tool)*, to give researchers the support they need to identify, assess, and address research participant risks in this fast-moving field.

Our experimental study found that use of the DHC-RM Tool led to a very large improvement in how well researchers managed the risks of digital health research studies. Using the toolkit, they were able to identify more risks than they identified using current practice–including risks they would not otherwise have considered. They were also able to come up with more changes to reduce the risks associated with digital health research studies, including changes they would not otherwise have considered. Those who used the toolkit found it beneficial and easy to use.

The DHC-RM Tool fills an important gap in the science and practice of participant protection in digital health research.

## Introduction

As digital health technologies (DHT) rapidly proliferate, researchers involving human participants in their studies find it increasingly challenging to select and use digital tools in ways that align with both scientific goals and ethical obligations. These DHTs, ranging from mobile apps and wearable sensors to artificial intelligence-based platforms, hold immense promise for enhancing data collection, improving participant engagement, and personalizing healthcare interventions [1]. However, the pace of innovation in the digital health landscape often outstrips the development of regulatory frameworks and best practices, leaving researchers without clear guidance on evaluating the scientific validity, ethical integrity, and practical utility of available technologies [2]. The process of selecting a DHT is further complicated by a lack of standardized criteria for assessing key dimensions such as data governance, participant privacy, user experience, and long-term feasibility. This gap has the potential to compromise both the quality of research outcomes and the safety of participants, particularly in studies involving vulnerable populations or novel, minimally tested digital tools.

In response to these challenges, Nebeker et al. (2020) [3] developed the *Digital Health Checklist for Researchers (DHC-R),* a structured decision-support tool designed to facilitate informed and ethically sound choices when integrating DHTs into research with human participants. The checklist synthesizes stakeholder input and ethical principles into an actionable framework that supports researchers in critically assessing DHTs across four key domains: access and usability, data management, risks and benefits, and privacy. By translating abstract ethical norms into tangible evaluation criteria, the DHC-R fills a critical gap in the research ecosystem, enabling investigators to navigate the complex digital health landscape with greater clarity and confidence.

The development of this checklist reflects a broader movement toward embedding ethics into the design and deployment of digital health research. As Nebeker et al. emphasize in a related publication, actionable ethics, particularly in studies involving artificial intelligence, must be integral to the research lifecycle rather than an afterthought [4]. Through its emphasis on interdisciplinary dialogue and contextual judgment, the DHC-R serves not only as a decision-making aid but also as a catalyst for promoting responsible innovation in digital health research. This checklist-based approach represents the current state of the art of risk management practice in research involving human participants [5–8]. Specifically, researchers and ethics review boards, such as institutional review boards (IRBs), often rely on checklists for identifying risks. These checklists ensure that stakeholders consider whether certain potential risks might apply to a research protocol. For instance, it is common to use a checklist that identifies whether a research project will include participants from vulnerable populations such as prisoners, children, pregnant women, etc. [9,10]. While checklists ensure that such risks are noted, they do not provide a structure for in-depth risk assessment (which also includes exploring the underlying causes of risks, their potential consequences, rating the likelihood and impacts of those consequences, and determining how much–if anything–should be done to modify those risks). And, to the extent that checklists promote action regarding these hazards, that action is typically limited to documentation and informed consent.

In the safety risk management literature (e.g. patient safety, occupational safety) risk identification is seen as a critical step in the risk management process, but one that is important only to the extent that it informs the later steps of risk analysis, risk evaluation, and risk control [11]. *Risk analysis* focuses on understanding the potential consequences that might result from a hazardous situation, how likely those consequences are to materialize, and how severe they might be. *Risk evaluation* uses the information developed through risk analysis to make decisions about whether (and to what degree) actions should be taken to reduce these risks. *Risk control* (also known as *risk treatment)* is the design and management of interventions to accomplish any risk reduction that is required [12–14].

In this study, we sought to advance the state of the art in protection of human participants for digital health research–first, by enabling more a more robust risk identification process that does not rely only on pre-specifying each individual risk of concern (an important consideration in this fast-moving field) and second, by providing more comprehensive support for the rest of the risk management process. To accomplish this, we developed and evaluated a novel Digital Health Checklist–Risk Management (DHC-RM) Tool.

## Materials and methods

### Materials

#### Design of the DHC-RM Tool

The DHC-RM Tool was developed using an iterative design process that included multiple rounds of internal review and five rounds of user feedback/revisions (in the form of cognitive walkthroughs [15] and user surveys) to arrive at a user-friendly interface that mimics a standalone DHC-RM Tool app. The tool was implemented via REDCap, a secure web platform for building and managing online databases and surveys [16].

The DHC-RM Tool combines the existing DHC with a complementary risk assessment tool, converting it from an approach that focuses solely on risk identification to one that supports comprehensive risk assessment for evaluating a digital health tool. It does this by abstracting key themes from the DHC and treating these as brainstorming prompts in a specialized version of the Structured What-If Technique (SWIFT) [13]. Most risk assessment tools use unstructured brainstorming to identify risks for assessment [13]. For instance, in a failure mode and effects analysis (FMEA), stakeholders first map out a process or system, and then simply ask, “what could go wrong?” for each step or component [17]. SWIFT, on the other hand, relies on *structured* brainstorming (brainstorming ways in which particular risk mechanisms might apply to the situation at hand) [13]. In a patient safety-focused SWIFT analysis, for instance, users might draw theory-based brainstorming prompts from the work systems component of the Systems Engineering Initiative for Patient Safety (SEIPS) 2.0 model (external environment, tools & technology, organization, tasks, internal environment, and persons) [18] and–one prompt at a time–brainstorm ways that each of those might give rise to harm.

The brainstorming prompts used in conducting a SWIFT analysis can be tailored to the situation at hand, which allows SWIFT to dovetail perfectly with the DHC. The iterative design process described above eventually distilled the themes from the DHC into 3 brainstorming prompts for the DHC-RM Tool: 1. **Access, usability, and fairness**; 2. **Data collection and management**; and 3. **Effectiveness, reliability, and safety.** In line with our prior work [12,19–21], each prompt is presented alongside just-in-time training to promote ease of use. This takes the form of a brief description of the terms and selected examples that demonstrate how the ideas can be applied. (See **Fig 1** for an example).

**Fig 1.**
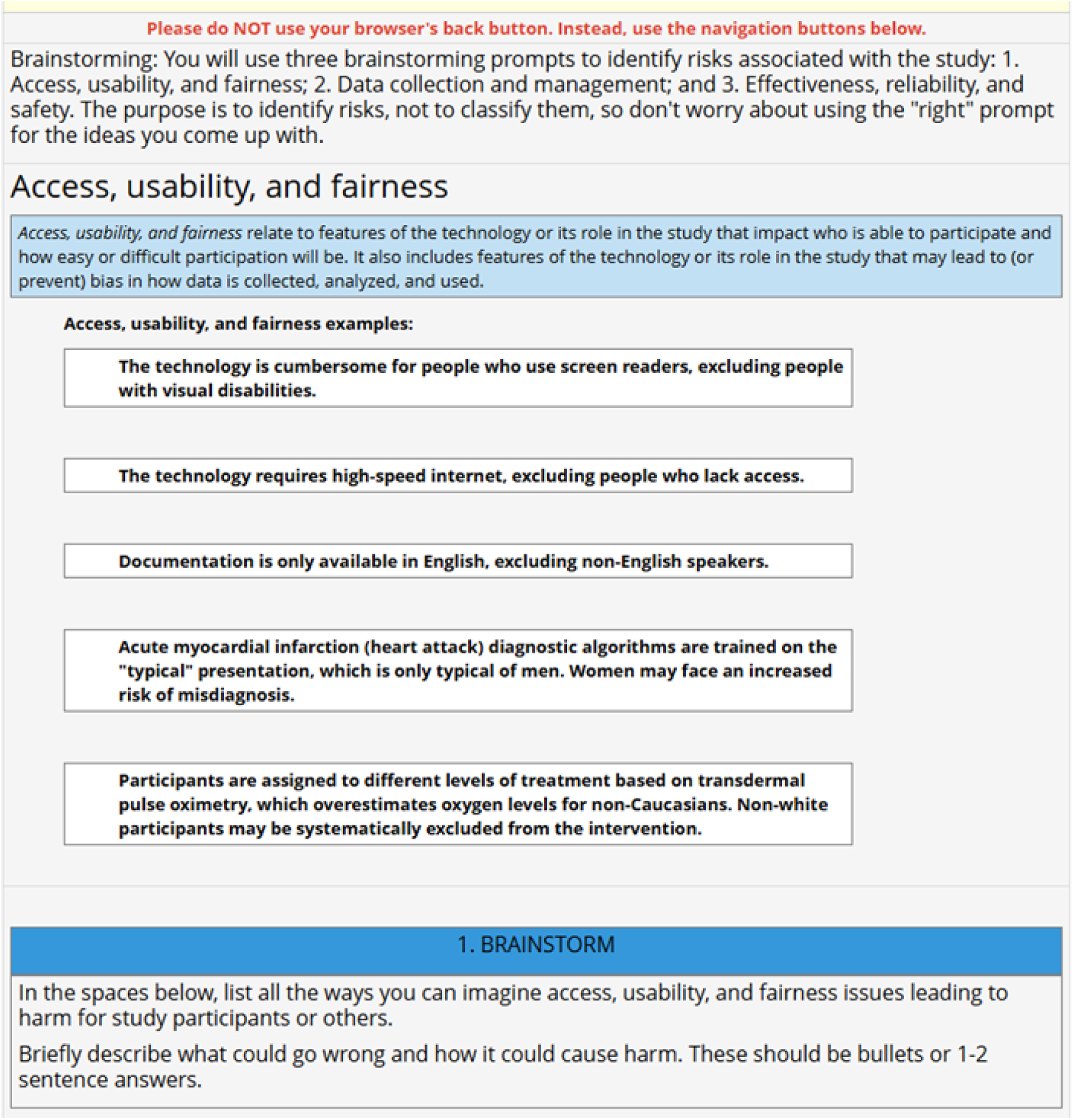
DHC-RM Web Prototype 0.2 - Access, Usability, and Fairness Brainstorming Prompt with description and examples.

Strictly speaking, SWIFT was developed as a risk identification tool, but in practice it is typically paired with a risk scoring procedure, a risk evaluation process based on the risk score, and a prompt to record risk control recommendations.(6) The DHC-RM Tool is intended for use in 1) improving participant protection in the design of research protocols, and 2) communicating risks, benefits, and protections associated with the research protocol to support ethics review. We therefore designed the tool to highlight the connection between identified risks, risk controls adopted in response to those risks, and the residual risk remaining in light of those risk controls–while also capturing the anticipated benefits of the study. The DHC-RM Tool designed to be a self-documenting process [12], in which working through the tool automatically produces a useful output for communicating the analysis and underlying rationales without further report-writing on the part of users. The high-level process for using the DHC-RM Tool is shown in **Fig 2** below.

**Fig 2.**
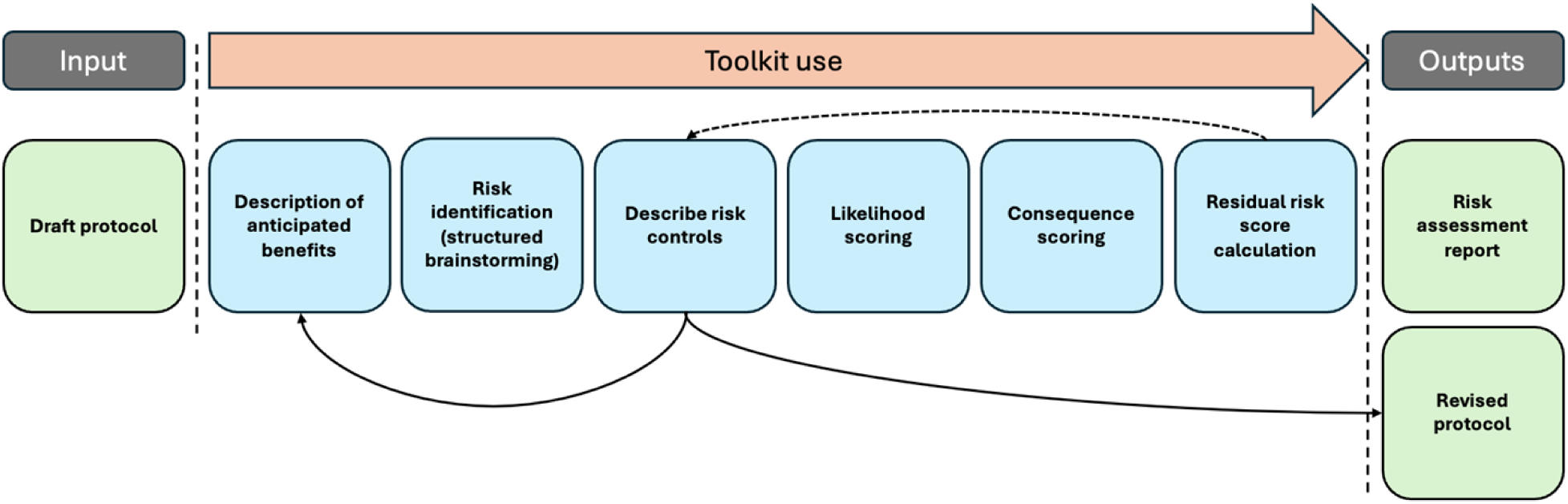
DHC-RM Tool Process.

Users of the toolkit are first asked to describe the anticipated benefits associated with the study under consideration. They next engage in a structured brainstorming process to identify risks, as described above. They then develop and record risk control interventions to reduce any risks that warrant action. Considering each risk in light of any risk controls that have been developed, users next rate the *likelihood* that the risk might occur on a scale of 1-4 and the *level of consequence* that would be expected if the risk did occur, also on a scale of 1-4. (The scoring guidance for these ratings is shown in **Table 1**.) The likelihood and consequence scores are automatically multiplied by one another to arrive at a final *risk score* for each identified risk (range 1-16). Risk scores are mapped onto one of 3 risk categories characterizing the overall level of residual risk represented. **High**, depicted in red (risk score ≥ 8), **medium**, depicted in yellow (4 ≤ risk score ≤ 7), or **low**, depicted in blue (risk score ≤ 3)]. If the residual risk remains unacceptably high, users should revisit the risk control stage and then rescore the risks to account for any changes. The tool automatically produces a concise risk report for communicating the results (and underlying rationales) to research ethics committee members and other stakeholders (e.g., a research student might use it to facilitate a discussion with their supervisor) and informs the development of a revised research protocol.

**Table 1.** Scoring guidance for likelihood and consequence.

| Scores | Likelihood | Consequence |
| --- | --- | --- |
| 4 | **Very high:** A more than 10 in 1,000 chance of occurring | **Very high:**<br>Impacts on individual wellbeing that may be life-altering (death, cure, long-term change in the ability to conduct activities of daily living, long-term change in social relationships, etc.)<br>Or severe societal impacts |
| 3 | **High:** More than a 5 in 1,000 chance of occurring, but less than a 10 in 1,000 chance of occurring | **High:**<br>Impacts on individual wellbeing that may be long-lasting but minor to moderate<br>Or temporary but major or moderate societal impacts. |
| 2 | **Low:** More than a 1 in 1,000 chance of occurring, but less than a 5 in 1,000 chance of occurring | **Low:**<br>Impacts on individual wellbeing that may be long-lasting but negligible<br>Or temporary and minor-to-moderate or permanent but minor societal impacts |
| 1 | **Very low:** A less than 1 in 1,000 chance of occurring | **Very low:**<br>Impacts on individual wellbeing that are temporary and negligible-to-minor |

## Methods

We conducted an online study with digital health researchers comparing the DHC-RM Tool to current practice. A convenience sampling strategy was used to recruit potential participants via professional networks, social media, and professional societies. Potential participants were also invited to share the recruitment message with others who might qualify. Eligible participants were adults, 18 years or older who were trained in digital health research design, and who had assisted in digital health research design, designed digital health studies under close supervision, or had supervised others in designing digital health research. Those who were eligible and consented to participate were then randomized into one of eight groups, as shown in Fig 6.

Participation included:

1. A brief demographic questionnaire
2. Using the DHC-RM Tool [See **Fig 3** for the experimental design]

a. Assessing the risks and benefits of one of two digital health research studies [**S1 Appendix or S2 Appendix**] using current practice (Phase 1)
b. Assessing the risks and benefits of one of two digital health research studies [**S3 Appendix** or **S4 Appendix**] using *either* current practice *or* the DHC-RM Tool (Phase 2)
3. A short feedback questionnaire (for those randomized to the DHC-RM Tool)

**Fig 3.**
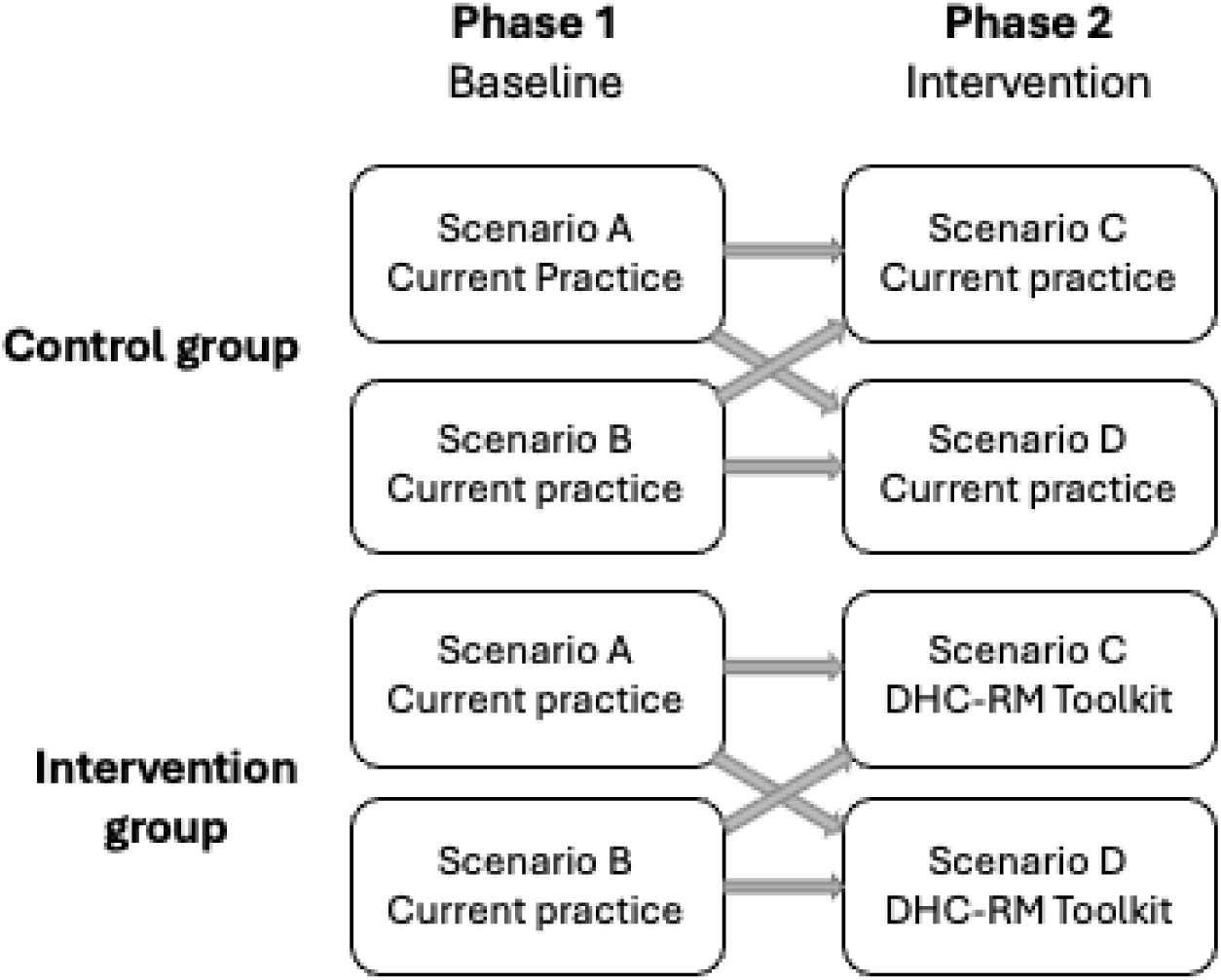
Experimental design.

To control for the potentially significant variation in baseline performance between participants, we used a randomized experimental difference-in-differences (REDiD) approach in line with our prior work [19]. Quantitative analysis focused on the intervention-related difference-in-differences in risks identified and risk controls proposed between phase 1 (baseline) and phase 2 (intervention).

We used evaluation metrics originally drawn from the design ideation literature (8) and previously used in the patient safety risk management literature (1): **A.** The quantity, novelty, and variety of risks identified (primary outcome measures), and **B.** The quantity, novelty, and variety of risk control / risk reduction strategies proposed to address identified risks (secondary outcome measures). **Quantity** is a measure of the overall count of on-topic risks identified or risk controls proposed. **Variety** relates to different overarching approaches (themes), rather than specific risks or risk control strategies (e.g., the theme of *data reporting*, vs. the specific strategy *only report data in aggregate*). **Novelty** is the degree to which specific risks/risk controls are “outside what would otherwise be expected” (1,8). Off-topic responses were not included in the main analysis. (For instance, in a scenario related to a study of people using methamphetamine, a risk identified as *potential overdose* was coded as off-topic because it pertained to the baseline risk in the study population, not to any additional risk imposed by the study.)

Quantitative data were developed through “small q” qualitative analysis (9) of risk identification and risk control outputs. Risks and risk control strategies were derived using qualitative content analysis (10,11). Risk identification coding was organized using six deductive themes (*access, usability, fairness, data collection & management, effectiveness & reliability,* and *safety*) with inductive coding of specific risks (e.g., *incidental collection of bystander data*). Risk control / mitigation strategies and related themes/subthemes were constructed using inductive coding alone. Codes were developed by a consensus of two researchers, with a third researcher available to resolve any differences. Codes were then converted to counts. Free-text qualitative feedback was also assessed through inductive coding. Statistical comparisons were made using two-sample t-tests.

### Data Collection

After randomization, participants were sent an initial personalized email within 24-48 hours of enrolling with instructions to complete the DHC-RM Tool survey in REDCap [16]. Reminder emails were scheduled to be sent every three days for up to three occurrences. Participants were given a two-week window to complete the survey. For surveys not completed by week two, a second personalized email was sent by the program coordinator. Surveys that were still not completed after a two-month period were closed and considered incomplete.

## Results

### Participants

In total, we recruited 72 digital health researchers who completed the initial screener and consented to participate. Of those 72, 63 were randomized, eight were dropped before randomization, and one refused. The eight dropped participants were identified as potential bots or scams. A total of 44 participants completed the study, but 4 were excluded from analysis due to erroneous responses (e.g., assessing the training examples rather than the scenario, or continuing to assess the Phase 1 scenario during Phase 2), resulting in a sample size of N=40 (control n=21, and intervention n=19; (See **Fig 4**).

**Fig 4.**
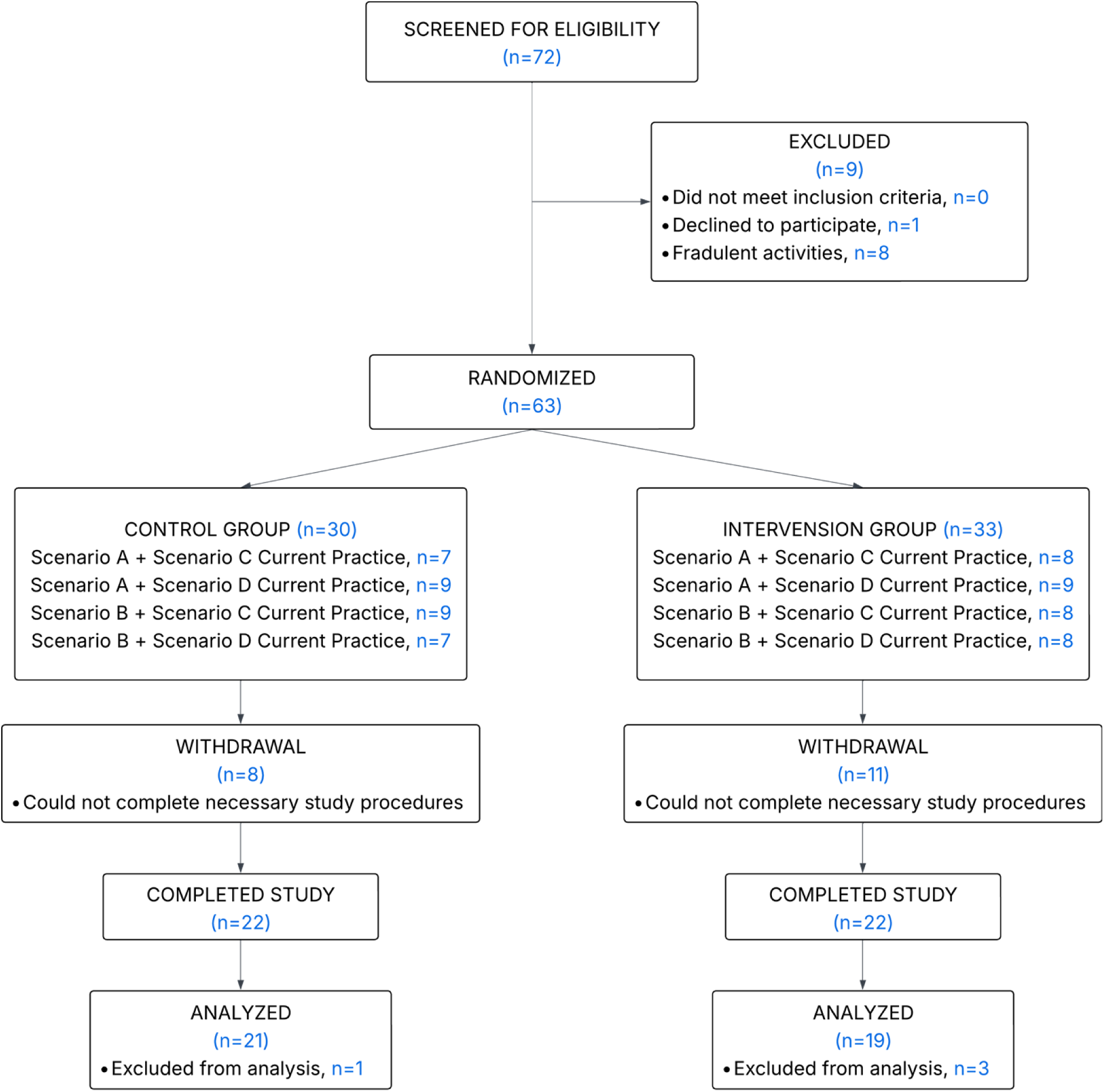
Recruitment Consortium Diagram.

### Risk identification

#### Phase 1 (Baseline)

Results were similar between the control and intervention groups at baseline, with no statistically significant difference in the overall number of risks identified per participant between the two groups. Nor were there statistically significant differences in the rate at which any specific themes or categories of risk were identified, although risks related to *fairness* were identified only by members of the control group (mean: 0.21 *fairness*-related risks identified per participant vs. 0 in the intervention group, p-value: 0.09). No participants from either group identified any risks related to *access* at baseline.

#### Quantity

Use of the DHC-RM Tool was associated with a large and statistically significant increase in the overall number of risks identified compared to baseline, with a mean of 14.7 additional risks identified per participant in Phase 2 for the intervention group vs. 0.26 for the control group. Table 2 shows the change in the total number of on-topic risks identified for each group in Phase 2 vs. baseline.

**Table 2.**
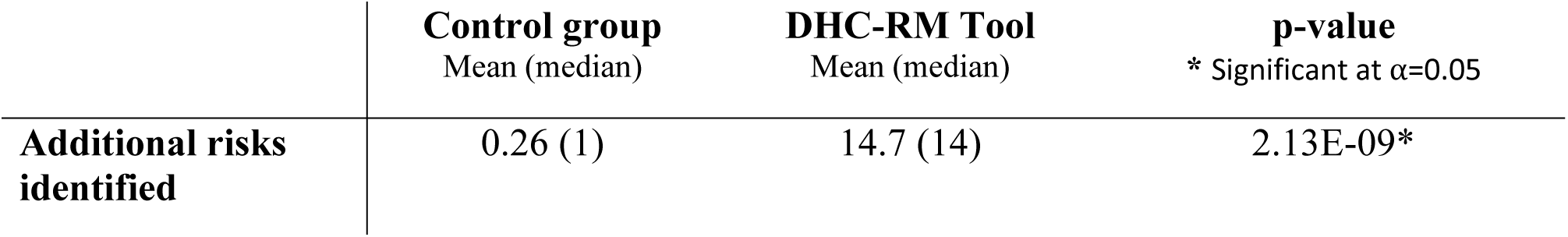
Difference in quantity of risks identified in Phase 2 vs. baseline by group.

#### Variety

##### Risk categories

We developed 6 risk identification themes for deductive coding:

###### Access

Risks in this category are characteristics of the DHT or study design that could exclude potential participants from taking part in digital health research, such as language barriers, lack of enabling technologies or services (e.g., smartphones, high-speed internet), or time constraints.

###### Usability

Risks in this category pertain to aspects of the DHT or study design that are not a bar to participation but make the study more onerous for participants. Usability barriers include problems with ease of use or user error, minor physical or psychological discomfort, and impaired ease of use for people with disabilities or other specific conditions.

###### Fairness

These risks relate to bias in data collection, bias in data analysis, or inequitable distribution of the risks and benefits of the digital health study.

###### Data collection and management

These risks involve an array of data-related issues, including the types and quantity of data collected, the means by which it is collected, data storage capacity, data security, data sharing, data ownership, loss of privacy and confidentiality, and incidental collection of bystander data (e.g., recordings of the participants that also capture family members who are not participants in the study).

###### Effectiveness and reliability

These risks relate to issues such as limitations and barriers inherent in the DHT, equipment malfunctions, unreliable results, and lack of explainability.

###### Safety

Safety risks include physical harms, psychological harms, legal harms, economic harms, social harms (other than legal/economic harms), societal harms (i.e., harm to society at a whole or to particular groups beyond the study participants), loss of autonomy, harm to bystanders, and non-specific harms (e.g., “Any possible identification of participants could be dangerous to individuals and their families”).

##### Variety in risk identification

Use of the DHC-RM Tool was associated with a practically and statistically significant increase in the average number of risks identified per theme across every theme, as shown in Table 3.

**Table 3.**
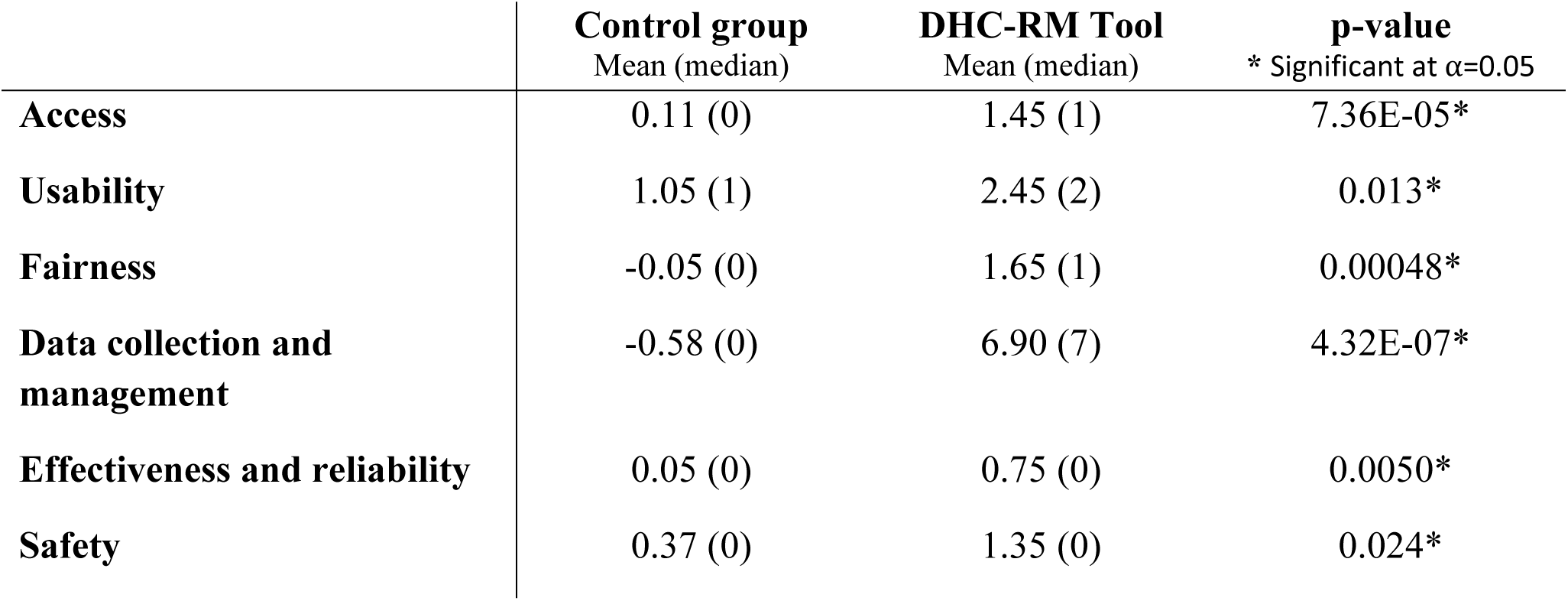
Impact on variety – Difference in the average number of risks identified by theme in Phase 2 vs. baseline.

Another way to consider variety is to evaluate, for each theme, the proportion of participants who identified at least one risk in Phase 2. Rather than attending to the *degree* of focus on each theme, as in the previous metric, this approach (in Table 4) evaluates the likelihood that participants will identify *any* risks in a given theme or miss that theme altogether.

**Table 4.** Impact on variety – Difference in the proportion of participants identifying ≥1 risk in a theme in Phase 2.

| | **Control group**<br>Proportion | **DHC-RM Tool**<br>Proportion | **p-value**<br>* Significant at $\alpha=0.05$ |
| --- | --- | --- | --- |
| **Access** | 0.11 | 0.75 | 4.94E-05* |
| **Usability** | 0.79 | 0.80 | 0.94 |
| **Fairness** | 0.16 | 0.60 | 0.0046* |
| **Data collection and management** | 1.0 | 0.95 | 0.32 |
| **Effectiveness and reliability** | 0.053 | 0.45 | 0.0045* |
| **Safety** | 0.37 | 0.49 | 0.15 |

Compared to the control group, users of the DHC-RM Tool were significantly more likely to identify risks related to *access, fairness, effectiveness and reliability*. There were no significant differences between the two groups for the themes of *usability, data collection and management,* or *safety.* Participants in the control group did not outperform DHC-RM Tool users in any theme.

#### Novelty

Use of the DHC-RM Tool was associated with much greater novelty in terms of the specific risks identified, as shown in Fig 5. Overall, 34 distinct risks were identified for Phase 2. Among these, 17 risks were identified exclusively by those using the DHC-RM Tool and 17 were identified by both approaches. None were identified exclusively by current practice. Not all digital health research scenarios involved the same risks, however, so we also considered scenario-specific novelty. A total of 29 distinct risks were identified for Scenario C: 19 by the tool alone, 10 by both approaches, and none by current practice alone. The results were similar for Scenario D, in which a total of 28 distinct risks were identified, with 14 identified exclusively by the DHC-RM Tool, 14 by both approaches, and none by current practice alone.

**Fig 5.**
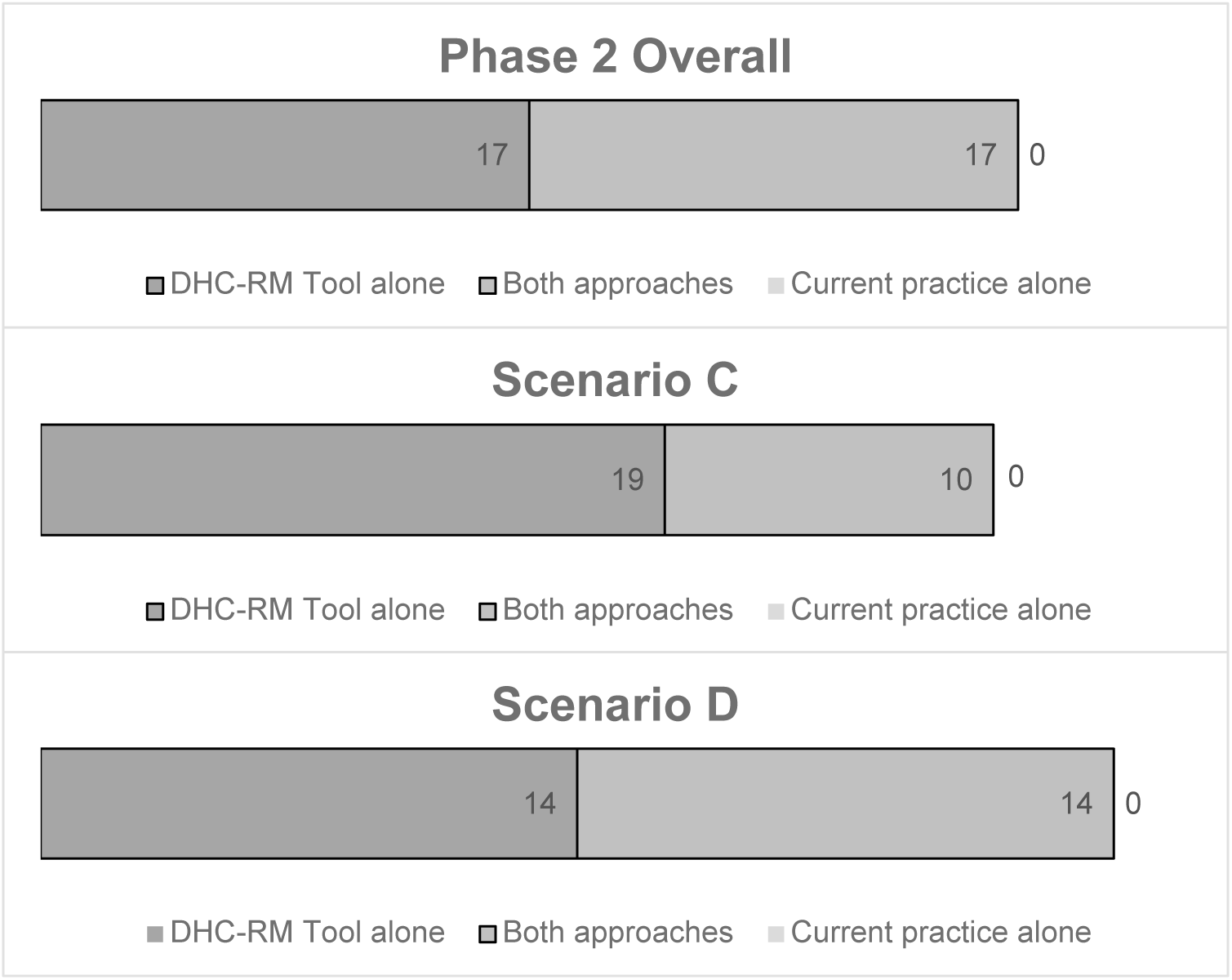
Novelty – Distinct risks identified.

The impact of the tool was statistically significant, as shown by Table 5, which evaluates the mean proportion of scenario-specific and pooled risks identified using each approach.

**Table 5.**
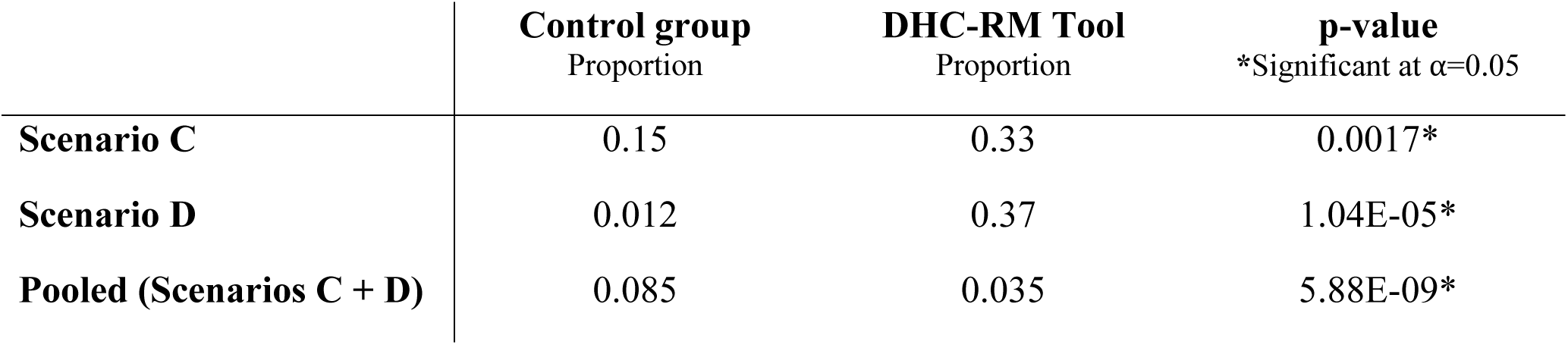
Scenario-specific novelty – Mean proportion of total scenario-specific and pooled risks identified by current practice vs. DHC-RM Tool.

### Risk control strategies

#### Quantity

Use of the DHC-RM Tool was associated with a large and statistically significant increase in the overall number of risk control strategies developed, with a mean of 9.63 additional risks controls produced in Phase 2 for the intervention group vs. 0.15 for the control group. Table 6 shows the change in total number of risk control strategies developed per participant for each group in Phase 2 vs. baseline.

**Table 6.** Difference in quantity of risk control strategies developed in Phase 2 vs. baseline by group.

| | **Control group**<br>Mean (median) | **DHC-RM Tool**<br>Mean (median) | **p-value**<br>* Significant at $\alpha=0.05$ |
| --- | --- | --- | --- |
| **Additional risk controls identified** | 0.15 (0) | 9.63 (10) | 4.2E-07* |

#### Variety

##### Risk control strategies

We developed 5 risk control themes and 6 sub-themes through inductive coding:

- Study design
- Informed consent
- Data management

- Data recording and processing
- Data security
- Data reporting
- Return of results
- Data sharing
- Participant responsibility for data protection
- Clinical action based on the study
- Adverse event management

##### Variety in risk control themes

Controlling for baseline, users of the DHC-RM Tool developed a higher average number of risk control strategies across all categories and subcategories. As shown in Table 7, these differences were statistically significant for *study design* (which showed the largest impact) and *data management*, as well as the subcategories *data recording and processing* and *data sharing*.

**Table 7.** Impact on variety – Difference in average risk control strategies developed for each theme/subtheme in Phase 2 vs. baseline.

| | **Control group**<br>Mean (median) | **DHC-RM Tool**<br>Mean (median) | **p-value**<br>* Significant at $\alpha=0.05$ |
| --- | --- | --- | --- |
| **Study design** | 0.7 (1) | 5.58 (6) | 3.98E-07* |
| **Informed consent** | 0.75 (1) | 1.37 (1) | 0.23 |
| **Data management** | -1.3 (0) | 2.58 (1) | 0.0050* |
| **Data recording and processing** | -0.85 (-0.5) | 0.32 (0) | 0.0048* |
| **Data security** | -0.4 (0) | 1.63 (0) | 0.091 |
| **Data reporting** | 0.05 (0) | 0.16 (0) | 0.51 |
| **Return of results** | 0 (0) | 0.16 (0) | 0.082 |
| **Data sharing** | -0.05 (0) | 0.26 (0) | 0.033* |
| **Participant responsibility** | -0.05 (0) | 0.053 (0) | 0.16 |
| **Clinical action** | 0 (0) | 0.053 (0) | 0.66 |
| **Adverse event management** | 0 (0) | 0.16 (0) | 0.45 |

*Subcategories of Data Management are shaded gray.*

As shown in Table 8, users of the DHC-RM Tool were significantly more likely to develop risk control strategies related to the theme of *study design,* and the subtheme of *data sharing*. There were no significant differences between the two groups for any other theme or subtheme. Of note, the subthemes of *return of results* and *data sharing* were developed exclusively by users of the tool.

**Table 8.** Impact on variety – Difference in in the proportion of participants developing ≥1 risk control strategy in a theme/subtheme in Phase 2.

| | **Control group**<br>Proportion | **DHC-RM Tool**<br>Proportion | **p-value**<br>* Significant at $\alpha=0.05$ |
| --- | --- | --- | --- |
| **Study design** | 0.6 | 1.0 | 0.0020* |
| **Informed consent** | 0.8 | 0.68 | 0.41 |
| **Data management** | 0.9 | 0.95 | 0.58 |
| **Data recording and processing** | 0.35 | 0.53 | 0.27 |
| **Data security** | 0.75 | 0.84 | 0.48 |
| **Data reporting** | 0.1 | 0.16 | 0.59 |
| **Return of results** | 0.0 | 0.16 | 0.064 |
| **Data sharing** | 0.0 | 0.21 | 0.030* |
| **Participant responsibility** | 0.05 | 0.053 | 0.97 |
| **Clinical action** | 0.15 | 0.053 | 0.32 |
| **Adverse event management** | 0.06 | 0.16 | 0.27 |

*Subthemes of Data Management are shaded gray.*

#### Novelty

Use of the DHC-RM Tool was associated with much greater novelty in terms of the risk control strategies developed, as shown in Fig 6. Overall, 110 risk control strategies were developed for Phase 2. Among these, 62 were developed exclusively using the DHC-RM Tool, 29 were developed using both approaches, and 19 were developed exclusively using current practice. Seventy-six distinct risk control strategies were developed for Scenario C: 51 by the tool alone, 12 by both approaches, and 13 by current practice alone. Scenario D also resulted in 76 distinct risk control strategies, of which 37 were developed exclusively by the tool, 19 by both approaches, and 20 exclusively by current practice.

**Figure 6.**
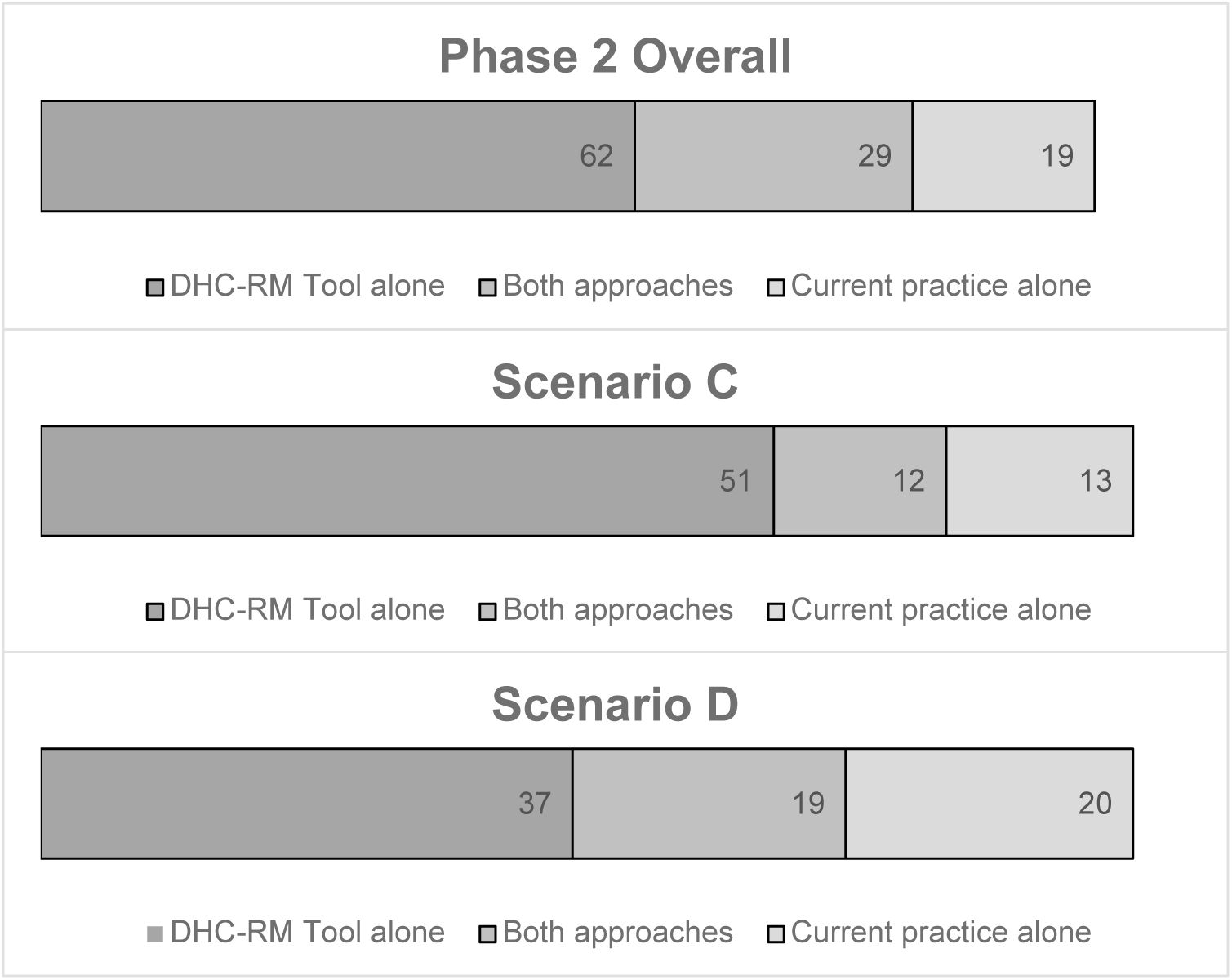
Novelty – Distinct risk control strategies developed.

The impact of the tool was statistically significant, as shown by Table 9, which evaluates the mean proportion of scenario-specific and pooled risk control strategies developed using each approach.

**Table 9.** Scenario-specific novelty – Mean proportion of total scenario-specific and pooled risk control strategies developed by current practice vs. DHC-RM Tool.

| | Control group<br>Proportion | DHC-RM Tool<br>Proportion | p-value<br>* Significant at $\alpha=0.05$ |
| --- | --- | --- | --- |
| **Scenario C** | 0.054 | 0.14 | 0.0017* |
| **Scenario D** | 0.074 | 0.13 | 0.042* |
| **Pooled (Scenarios C + D)** | 0.064 | 0.13 | 0.00010* |

### User feedback

We collected feedback from participants in the intervention group (those who used the current practice in Phase 1 and the DHC-RM Tool in Phase 2) (n=20).

#### Willingness to use

Fifteen participants (75%) said they would use the tool again. Among those who would use the tool again, common reasons included: *a structured, systematic approach that increases confidence in participant protections* (e.g., “Systemizing information of the evaluation of risks and benefits provides a structured approach to this assessment that makes me feel more comfortable in providing and ensuring participant safety”); that it *generated new insights into risks* (e.g., “Oftentimes, it may be difficult to think of the risks off the top of your head. Brainstorming and having a different visual representation of possible issues and how to mitigate them [allows] for a more holistic perspective.”); *useful examples* that “are helpful to get the juices flowing;” the *color-coded presentation of risk scores* (e.g., “The tool, by color coding, identifies high risks which I did not realize the amount of risk until the summary page. Indeed, [I] want to go back for more thoughts on risk mitigation…”); and *ease of use* (e.g., “Instructions are precise and topical to the specific step in the DHC-RM tool (not drinking through a firehose)).”

Less common themes included the fact that it is based on “an established, proven framework;” that it enables more holistic analysis; that it can be used collaboratively to integrate insights across the research team (e.g., “Have individuals provide information required by the tool to get different viewpoints. Compile the information to come out with one assessment that could be presented to a pre-IRB review…. getting the clinical team to provide their assessment would help the [team] come to consensus on mitigations along with accepting some of the higher level risks.”); that it can be used for communicating risks and risk reduction strategies to key stakeholders (e.g., “Providing this information to IRB Onboarding for new people coming into the study to help them review risks and mitigation.”); that it reminds users of ethical concerns; and that it meets standards of practice from other safety-critical healthcare research and development domains (e.g., “I came from the medical device world, where this type of risk assessment is built into the process through FMEA, and when I got into drug development, was surprised to learn that a similar type of tool/exercise is not commonly used. This is very helpful to the team to focus attention on risk assessment and mitigation.”).

Among those who would not use the tool again, the only theme mentioned by more than one participant was a lack of clarity about how to interpret risk scores (e.g., “What does a high risk score *mean*?”). Themes mentioned by only one participant included: Difficulty quantifying risks; potential subjectivity in risk scores; the tool being “a little complex to use;” that it was sometimes hard to know how to categorize risks (e.g., “For example, drawing lines between accessibility/usability/data collection and analysis can at times be ambiguous and difficult to categorize.”) [Note: Users are not asked to categorize risks as part of the DHC-RM Tool process.]; and that the participant is “more accustomed to deferring to the knowledge and experience of the PIs and research coordinators on the studies I work on.” One participant noted that they “…would be willing to use a future version of the tool, if it was further refined.”

#### Suggested improvements

We also asked how the DHC-RM Tool could be improved. Two participants said that *no improvements were required*. This included one respondent who said they would not use the tool again (because they would, instead, defer to the expertise of the PI or research coordinators). The most common suggestion was for *additional examples*. This included requests for more examples in general, for example sets tailored to specific scenarios (e.g., “The ability to request examples that are better correlated with the study being evaluated [e.g. social science, psychology, medical devices, therapeutics, etc.]”), and for examples of risk reduction strategies (e.g., “I wish there were also examples of strategies to respond to common risks. I had trouble coming up with strategies for mitigation.”). Three participants suggested *a checklist-based approach* (e.g., “Provide a long checklist of potential harms and possible mitigation strategies, to help with brainstorming and to not re-invent the wheel with every application”). *Additional instructions (and instructions at a lower reading level)* were also suggested by more than one participant (e.g., “Lower reading level/ shorter sentences for instructions. Although instructions are precise and topical to the specific step in the DHC-RM tool (not drinking through a firehose) -- I still had to re-read many times to feel confident that I understood what was being asked of me.”) Suggestions made by only one participant included: Making the tool *“more dynamic”* (not further specified); *integrating a large language model* to assist with generating risk reduction strategies; *reducing the length* of the tool; restructuring the tool to *focus on one risk at a time*; involve *more than one person* when using the tool; *limiting the number of entries in each category* to force users to refine their thinking; and *better ordering of examples* (not further specified).

#### Benefits of use

We also asked participants about their agreement with several questions related to the potential benefits of using the DHC-RM Tool. Responses were scored on a scale of 1-5. As shown in Table 10. The median score for all questions was a 4.00, showing good overall agreement with the anticipated benefits of adoption. Nineteen participants responded to this portion of the survey, one of whom skipped one question.

**Table 10.** Agreement with the potential benefits of the DHC-RM Tool.

|  | **N** |  | **Mean**<br>**(median)** | **Std.**<br>**Deviation** |
| --- | --- | --- | --- | --- |
| **Statement** | **Valid** | **Missing** |  |  |
| **The DHC-RM Tool is easy to use.** | 18 | 1 | 3.83 (4.00) | 1.098 |
| **Using the DHC-RM Tool would make it easier to show the IRB that I have considered all the risks associated with my digital health research protocols.** | 19 | 0 | 4.05 (4.00) | 1.026 |
| **This process inspired me to think of new potential harms of the research.** | 19 | 0 | 4.05 (4.00) | 1.079 |
| **This process inspired me to think of new risk reduction strategies.** | 19 | 0 | 3.74 (4.00) | 1.24 |
| **I will feel more confident in the safety of my digital health studies if I use the DHC-RM Tool.** | 19 | 0 | 3.63 (4.00) | 1.065 |
| **I think my IRB should encourage digital health researchers to use the DHC-RM Tool.** | 19 | 0 | 3.89 (4.00) | 1.15 |
| **Using the DHC-RM Tool would probably reduce the number of revisions requested by the IRB when I submit a digital health protocol.** | 19 | 0 | 3.58 (4.00) | 1.071 |
| **I would recommend using the DHC-RM Tool to other digital health researchers.** | 19 | 0 | 3.84 (4.00) | 0.958 |

## Discussion

The theory-based DHC-RM Tool drastically improves risk management practice for participant protection in digital health research–with impacts that are not only statistically significant, but also practically significant (e.g., compared to current practice, the increased number of risks identified and risk controls developed were more than 56 times as high and 64 times as high, respectively). By demonstrating that the research community easily *can* do better, this study shows that we *must* do better. As with the analogous work of patient safety [22], we cannot sit content in our good intentions and our “standard of practice” for human participants protection–especially when faced with the pervasive and often poorly understood risks of digital health research [23].

In risk management, you cannot manage what you cannot imagine. Some of the risks inherent in digital health research are immediately obvious; others are not [23–27] and unimagined harms go unaddressed. Digital health research teams often lack formal training in key domains of risk identification: Behavioral and clinical scientists report limited preparation for handling the ethical, legal, and social implications of digital tools, while technology developers frequently lack expertise in privacy and security best practices [28,29]. Analyses of mobile and connected health technologies demonstrate that important risks are routinely under-addressed in practice (including opaque data sharing, secondary use of data, and inequitable impacts on marginalized groups), suggesting systematic gaps in the way risks are identified and managed [30–33]. Our findings show the Digital Health Checklist–Risk Management (DHC-RM) Tool can help overcome these deficits by systematically expanding the risk landscape that digital health researchers consider.

Merely identifying risks is not enough, however. To bring participant protection in line with other safety risk management fields, researchers also need support for risk evaluation and risk control. All studies have *some* risk, however minimal, and ethical protection of research participants relies on balancing these risks against anticipated benefits. The risk evaluation / risk report component of the tool provides a structured and systematic approach to considering (and, importantly, *communicating*) the magnitude of the risks involved in a digital health study. A high residual risk score may also prompt researchers to develop additional risk controls (or rethink the study design from scratch) before any harm can occur. It is worth noting that there is no widely accepted standard for risk scoring guidance. The scoring cutoffs used in this study (Table 1) could be adapted to meet local needs.

Even in fields with a much longer tradition of using formal risk management tools, such as patient safety and occupational safety, risk control design remains a challenge [34–36]. The current standard of practice in these fields is, as embodied in the DHC-RM Tool, simply to prompt users to record risk controls in response to unacceptable risks. In contrast with the risk assessment process, there is no direct support for the actual generation of design concepts or vetting of potential risk controls [13,22,34,37,38]. There is evidence that tools to provide such support can improve the risk control process, however [12,19–21,39,40], and in future work we will consider whether they should be incorporated into the next generation of the DHC-RM Tool. Meanwhile, our findings show significant benefit to risk control practice when it is informed by stronger risk assessment and a structured process to help connect the dots between identified risks and proposed risk controls.

The tool appears to have a particularly strong impact on risk controls related to foundational aspects of study design, as well as issues frequently overlooked in digital health research, such as return of results to study participants and data sharing practices. Improved risk control practice related to these upstream and downstream issues suggest the DHC-RM Tool can help broaden the solution space that researchers engage with and help tackle issues related to invisible or secondary uses of data. The structure of the tool also helped promote good risk control coverage of identified risks and a clear and logical correspondence between risk controls and the risks they were meant to address–the lack of which remains a problem in safety risk control practice more broadly [41][42].

The overwhelmingly positive user feedback we received suggests that researchers would be willing to use the tool and recommend its use by others. This is important because the impact of any practice tool is the product of both efficacy *and implementation*. In the absence of regulatory or administrative requirements, uptake of the tool will only proceed if researchers see it as valuable (i.e., perceive that the *benefits* of use, such as improved participant safety and fewer rounds of ethics review, outweigh the *sacrifices* associated with use, such as a modest additional time investment) [43,44]. Participants also provided useful suggestions for the next iteration of the tool, including more examples / tailorable examples, providing additional support for the risk control section, and clarifying some concepts. We were able to draw implicit guidance for improvement from participant responses, as well: For instance, one user was concerned with classifying risks correctly in the risk identification stage, which is not an intended function of the tool. (To paraphrase our prior work, *what matters is that all categories of risk are considered, not which “bucket” a particular risk is placed in).* [45]

### Other potential applications of the DHC-RM Tool

Although the DHC-RM Tool was primarily designed to guide the work of researchers developing research protocols and IRB members reviewing them, it may also be useful in other contexts. For instance, technology developers could use the tool to gain insight into the potential risks associated with their design decisions. Few technologists are cross-trained in the ethics, health, and social science domains that inform the tool, and “current [technology] design methods are rarely helpful in guiding designers to incorporate human values.” [46] The DHC-RM Tool can help bridge this gap. It could also be used to facilitate genuine (rather than tokenistic) [47,48] patient/stakeholder engagement in research studies by providing a structured way to elicit insights. More effective stakeholder engagement from those whose lived experience could provide greater insight into the real-world risks and benefits of study design decisions. The DHC-RM Tool could also be useful for ongoing risk monitoring as part of longer-term research projects [49]. Outside of the research domain, the tool could inform decisions about whether–and how–to incorporate DHTs into routine patient care and occupational settings.

### Conclusion

Managing risks to participants in the quickly-evolving field of digital health research is not easy– but it is important. Just as in healthcare practice [50], there is an ethical duty to pursue excellence and effectiveness, rather than settling for compliance and complacency. The current standard of practice in human research protections, however, lags well behind safety risk management practice in other fields. This can lead to harm for study participants, bystanders, the research enterprise, and society as a whole. The DHC-RM Tool bridges this gap by introducing state-of-the art risk management tools for assessing and addressing risks in the design of digital health studies. It significantly improves practice–and thus the safe design of digital health research.

## Data Availability

The data underlying our study findings are available upon request to the corresponding authors.

## Acknowledgements

We would like to acknowledge the insights of our Study Advisory Committee (SAC) Members, Drs. Rebecca Ellis and John Torous, for their supportive and engaged involvement during the project.

For data collection, the REDCap software system was provided by the UCSD Clinical and Translational Research Center supported by Award Number UM1TR005449 from the National Center for Research Resources.

Research reported in this article was funded through a Patient-Centered Outcomes Research Institute® (PCORI®) Award (ME-2020C3-21310). The statements presented in this article are solely the responsibility of the author(s) and do not necessarily represent the views of the PCORI®.

## Supporting Information Captions

**S1 Appendix.** Study Scenario A

**S2 Appendix.** Study Scenario B

**S3 Appendix.** Study Scenario C

**S4 Appendix.** Study Scenario D

